# Stage-Specific Associations of Automated RPR With *Treponema pallidum* qPCR Positivity and Cycle Threshold in Early Symptomatic Syphilis: A Multicenter Study Using Specimens Collected During Routine Care

**DOI:** 10.64898/2026.10.14.26365521

**Authors:** Ayu Kasamatsu, Yuki Ohama, Yui Tomo, Kazuo Imai, Shu-ichi Nakayama, Ryosuke Omori, Takuya Yamagishi, Mitsuru Yasuda, Yukihiro Akeda

## Abstract

**Background:** *Treponema pallidum* quantitative polymerase chain reaction (qPCR) positivity and cycle threshold (Ct) by rapid plasma reagin (RPR), disease stage, and other characteristics are poorly characterized in routine-care specimens, leaving it unclear when qPCR of such specimens is informative for diagnosis.

**Methods:** This Japanese multicenter cross-sectional study explored qPCR positivity and Ct by these characteristics. Nonblood specimens collected during routine care underwent qPCR at a central laboratory for this study. Bayesian logistic and normal regressions modeled positivity and mean Ct of *TP0574* and *polA* with stage-specific continuous automated RPR terms, patient characteristics, collection site, and facility random intercepts. Estimates were standardized over the analyzed specimens. Population-group predictions were instead averaged within each group.

**Results:** Among 154 nonblood specimens from 145 patients (71.7% non-MSM men and women) at 14 facilities, 109 (70.8%) were qPCR positive. At RPR 1, adjusted positivity was 72.2% (95% credible interval, 60.9%–82.9%) in primary and 43.8% (16.7%–69.9%) in secondary syphilis. Positivity showed a modest downward trend with RPR in primary but increased in secondary syphilis, reflecting a stage-by-RPR interaction (secondary-to-primary ratio of odds ratios, 1.78; 1.25–2.65). Predicted positivity showed an upward trend with RPR values among MSM; adjusted positivity was relatively high for genital/urethral/urinary and anal/perianal/rectal specimens. Adjusted mean Ct increased with RPR in primary and followed an inverted-U pattern in secondary syphilis.

**Conclusions:** In primary syphilis, nonblood qPCR positivity was high even at low RPR. When serologic findings remain inconclusive in this stage, qPCR is highly recommended for securing an early diagnosis.

**Summary:** In nonblood specimens collected during routine care, *Treponema pallidum* qPCR positivity was high even at low automated RPR values in primary syphilis, potentially supporting earlier diagnostic confirmation when serology is inconclusive. In secondary syphilis, positivity increased with RPR.

## INTRODUCTION

Syphilis has resurged globally [1], with marked increases among heterosexual men and women in the United States [2] and Japan [3, 4]. Primary and secondary syphilis, the early symptomatic stages, contribute substantially to further spread. Accurate diagnosis at these stages is therefore central to patient care and to interrupting transmission.

Serologic testing establishes most diagnoses but may be inconclusive when rapid plasma reagin (RPR) values are low, before the nontreponemal antibody response has fully developed. In such cases, quantitative polymerase chain reaction (qPCR) can support the diagnosis by directly detecting *Treponema pallidum* (*T. pallidum*) DNA. When RPR values are high and serology is diagnostic, qPCR may still help determine whether a local manifestation of uncertain cause involves *T. pallidum* [5]. In both settings, it is useful for clinicians to know how likely qPCR is to be positive given RPR, disease stage, and other patient and specimen characteristics.

Recent studies have estimated qPCR positivity in relation to such characteristics using protocol-specified sampling from predefined anatomical sites [6–9]. In routine care, however, treating physicians choose sampling sites according to the presenting manifestations, and collection follows local practice. These differences may limit the applicability of existing estimates and leave it unclear when qPCR of clinically selected specimens is informative for diagnosis. Moreover, several of these studies predominantly enrolled men who have sex with men (MSM) [6–8], although stage and lesion distribution, which may influence positivity, differ among MSM, non-MSM, and women [10, 11].

Beyond positivity, qPCR provides cycle threshold (Ct) values that reflect relative treponemal DNA recovery at the sampled site and may bear on transmission potential [7–9], again with few data outside protocol-based studies. qPCR has also been applied to blood, which is already collected for serologic testing, but reported positivity varies across studies and methods [5, 8, 12].

We therefore aimed to estimate qPCR positivity and Ct in early symptomatic syphilis, in relation to automated RPR, syphilis stage, and other patient and specimen characteristics, using nonblood specimens collected during routine care at multiple Japanese facilities. We also aimed to estimate blood qPCR positivity in this population.

## METHODS

### Study Design and Setting

We conducted an exploratory multicenter cross-sectional study using specimens submitted from 28 facilities across 15 prefectures in eastern, central, and western Japan between November 2023 and March 2026. The submitting facilities ranged from sexual-health, dermatology, urology, and internal medicine clinics to general hospitals. Physicians were invited to submit specimens collected during clinical evaluation from patients with suspected or diagnosed syphilis to the central laboratory at the National Institute of Infectious Diseases (NIID), together with a case report form. Syphilis was diagnosed with reference to the Japanese Society for Sexually Transmitted Infections guideline [13], which requires reactive treponemal and nontreponemal serologic tests, a positive nucleic acid amplification test, or both. Nucleic acid amplification testing for *T. pallidum* is not reimbursed in Japan and is performed mainly at the NIID and prefectural public health institutes. For this study, the central laboratory performed *T. pallidum* qPCR on all submitted specimens and returned the results to the submitting physician.

### Specimen and Patient Selection

Patients diagnosed with primary or secondary syphilis were eligible for analysis. This included patients with low RPR values who were diagnosed by qPCR despite insufficient serologic evidence. The main analysis of qPCR positivity and mean Ct included all specimens from eligible patients except blood and central nervous system specimens. An additional analysis included blood specimens with available qPCR results from eligible patients.

### Clinical Variables

The case report form was completed by the treating physician at specimen submission and updated when additional information became available. It recorded syphilis diagnosis and stage, specimen material and collection site, age, sex assigned at birth, symptoms, sexual-contact history, previous syphilis, human immunodeficiency virus (HIV) infection status, and serologic results, including automated RPR values measured at the submitting facility or by a commercial laboratory. These assays, which are commonly used in Japan, report quantitative values rather than doubling dilutions [14]. Treating physicians assigned syphilis stage using the notification criteria of Japan’s National Epidemiological Surveillance of Infectious Diseases [15]. They selected the material and collection site at their discretion. Population group (MSM, non-MSM men, or women) was defined by sex and sexual-contact history.

### Outcomes

A specimen was considered qPCR positive if both targets, *TP0574* (TpN47) and *polA*, yielded a Ct value, or if one target yielded a Ct value below 36 that was reproduced by repeat testing of the same DNA extract [16]. Mean dual-target Ct was calculated by averaging the two target-specific Ct values among specimens positive for both targets.

### Specimen Processing and Dual-Target qPCR

Specimens were stored at 2–8°C after receipt until DNA extraction. Swabs were suspended in Tris-EDTA buffer, and DNA was extracted using the DNeasy Blood & Tissue Kit (QIAGEN, Hilden, Germany) following the manufacturer’s instructions.

Each DNA extract was tested using two singleplex hydrolysis-probe qPCR assays targeting *TP0574* (TpN47) [17] and the DNA polymerase I gene *polA* [16]. Amplification and fluorescence detection were performed using a LightCycler 96 System (Roche Diagnostics, Mannheim, Germany). All reactions were performed using TaqMan Gene Expression Master Mix (Applied Biosystems, Thermo Fisher Scientific, Waltham, MA, USA). Each run included positive, no-template, and internal amplification controls. Lower Ct values were interpreted as indicating greater recovery of treponemal DNA.

In November 2025, the *TP0574* primers and probe were replaced with those described by Romeis et al. [18], and amplification cycles for both assays were concurrently reduced from 45 to 40. The *polA* primers and probe remained unchanged throughout the study period. Additional details of blood specimen-processing are provided in the Supplementary Methods.

### Statistical Analysis

Categorical variables were summarized as counts (percentages) and continuous variables as medians (interquartile ranges). Observed qPCR positivity was tabulated by automated RPR category. The Pearson correlation coefficient between the two target-specific Ct values was calculated.

qPCR positivity and mean Ct were modeled using Bayesian logistic and normal regression, respectively. Both models included automated RPR, collection site, stage, and a facility random intercept, with adjustment for age, sex, HIV infection status, and previous syphilis history. For automated RPR, we used a log_2_(RPR + 1) transformation, with a linear term and its interaction with stage in the positivity model and stage-specific linear and quadratic terms in the mean-Ct model. Neither model included a patient-level random intercept because few patients contributed multiple specimens. Missing age and RPR values were estimated jointly with the regression parameters. Regression coefficients and variance components were assigned weakly informative priors.

Using posterior draws from each model, we estimated adjusted qPCR positivity and mean Ct by stage and collection site and across RPR values for each stage and for both stages combined, standardizing over specimens in each analysis while retaining the draw-specific values of facility effects. Population-group qPCR predictions were instead averaged within each group, preserving its covariate and facility composition; group-specific mean Ct was not estimated because some groups had few qPCR-positive specimens. Stage-specific adjusted odds ratios (aORs) per doubling of (RPR + 1) were obtained by exponentiating the posterior samples of corresponding coefficients, and their secondary-to-primary ratio quantified the stage-by-RPR interaction. For these exploratory analyses, posterior medians and equal-tailed 95% credible intervals (CrIs) were reported.

Posterior sampling used Hamiltonian Monte Carlo. Standard diagnostics [19] assessed convergence and sampler performance, and posterior predictive checks assessed model fit. Linear and quadratic RPR specifications were compared using specimen-level Pareto-smoothed importance-sampling leave-one-out cross-validation [20].

For both outcomes, sensitivity analyses made one change at a time: excluding patients with previous syphilis, restricting analyses to patients with recorded age and RPR, using narrower priors for selected outcome-model coefficients, or defining the outcomes using only *polA*, whose primers were unchanged. An additional qPCR-positivity analysis excluded patients for whom the qPCR result may have influenced the recorded diagnosis and thus eligibility. For mean Ct, an additional analysis used narrower priors for the stage-specific quadratic coefficients. Further statistical details are provided in the Supplementary Methods.

Finally, qPCR positivity in blood specimens was estimated overall and by syphilis stage, using Clopper–Pearson 2-sided 95% confidence intervals (CIs). All analyses were performed using R 4.5.3 with RStan 2.32.7 (Stan 2.32.2).

### Ethics

This study, including the opt-out consent procedure, was conducted in accordance with the Declaration of Helsinki and approved by the Ethics Review Committee of the NIID (approval no. 1605). Specimens and data were transferred without direct identifiers, and no key linking specimens to patients was provided to the NIID.

## RESULTS

### Specimens and Patients

The specimen-level analyses included 154 nonblood specimens from 145 patients with early symptomatic syphilis (primary, 108/145 [74.5%]; secondary, 37/145 [25.5%]) across 14 facilities. Of these, 109 (70.8%) were qPCR positive and 107 yielded Ct values for both targets (Figure 1). Non-MSM men and women comprised 71.7% of patients (Table 1). Median automated RPR (interquartile range [IQR]) was 7.3 (0.5–28.2) in primary and 88.9 (15.8–222.1) in secondary syphilis. Other characteristics by stage and specimen counts by assay configuration are summarized in Supplementary Tables S1 and S2, respectively.

**Figure 1.**
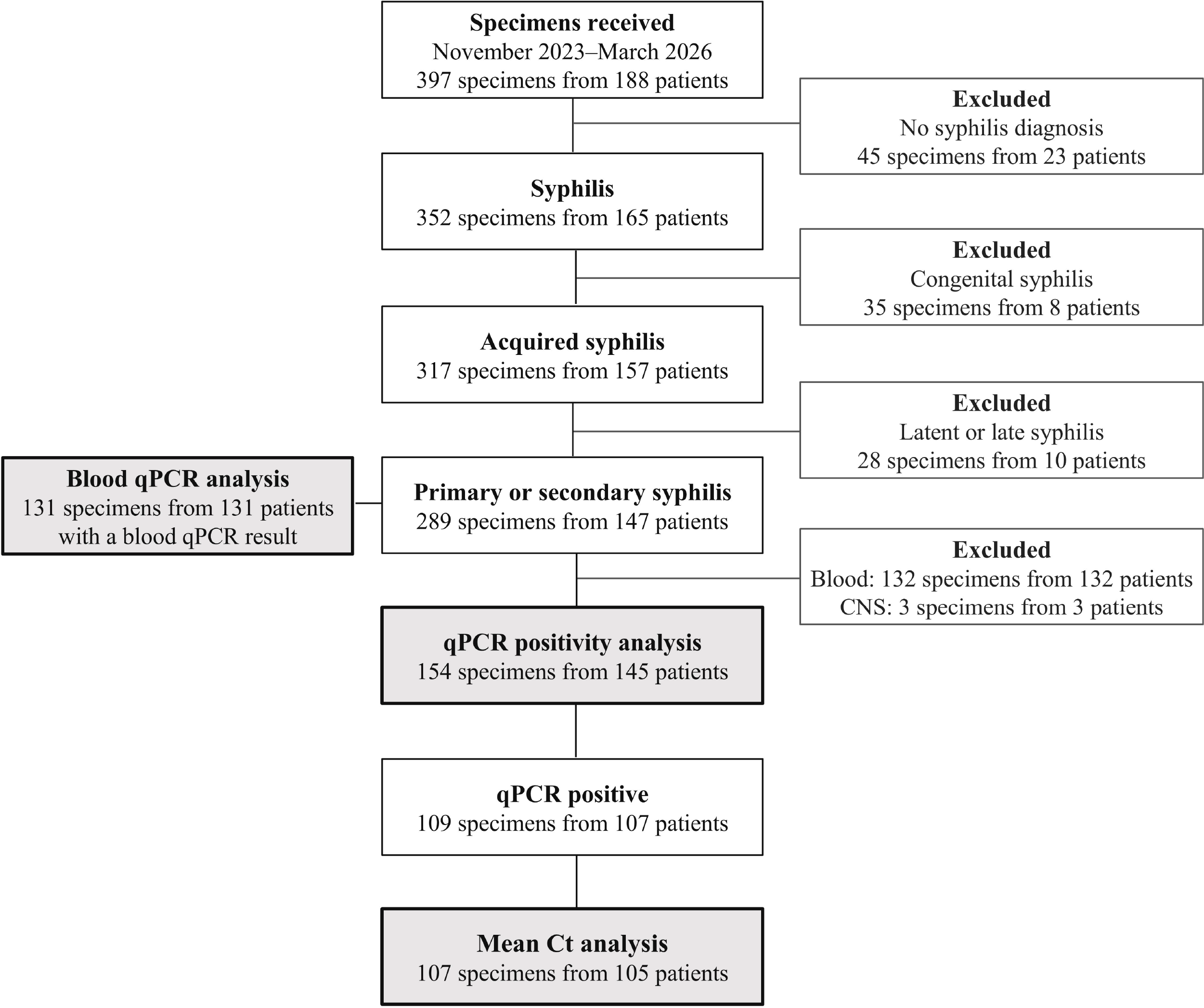
Selection of Specimens and Patients for Analyses. Numbers are specimens and unique patients unless otherwise indicated. Patient counts across specimen-based branches are not additive because patients could contribute to multiple specimen types. Abbreviations: CNS, central nervous system; Ct, cycle threshold; qPCR, quantitative polymerase chain reaction.

**Table 1.** Characteristics of Patients and Specimens.

A. Patient-level characteristics
| Characteristic | All patients<br>(*n* = 145) | qPCR-positive<br>patients<br>(*n* = 107) | Patients included in<br>mean Ct analysis<br>(*n* = 105) |
| --- | --- | --- | --- |
| Age, y, median (IQR) | 41.0 (32.5–51.0) | 41.0 (33.0–51.0) | 41.0 (32.5–51.0) |
| Age missing | 6 | 2 | 2 |
| Syphilis stage |  |  |  |
| Primary | 108 (74.5) | 83 (77.6) | 82 (78.1) |
| Secondary | 37 (25.5) | 24 (22.4) | 23 (21.9) |
| Population group |  |  |  |
| MSM | 41 (28.3) | 26 (24.3) | 25 (23.8) |
| Non-MSM men | 95 (65.5) | 74 (69.2) | 73 (69.5) |
| Women | 9 (6.2) | 7 (6.5) | 7 (6.7) |
| HIV infection | 22 (15.2) | 14 (13.1) | 13 (12.4) |
| Previous syphilis | 31 (21.4) | 16 (15.0) | 15 (14.3) |
| Automated RPR, median (IQR) | 12.0 (0.9–63.8) | 11.9 (0.9–40.8) | 11.6 (0.9–40.3) |
| Automated RPR missing | 11 | 5 | 5 |

B. Specimen-level characteristics
| Characteristic | All specimens<br>(*n* = 154) | qPCR-positive<br>specimens<br>(*n* = 109) | Dual-target-positive<br>specimens<br>(*n* = 107) |
| --- | --- | --- | --- |
| Collection site |  |  |  |
| Genital/urethral/urinary | 122 (79.2) | 95 (87.2) | 94 (87.9) |
| Anal/perianal/rectal | 17 (11.0) | 9 (8.3) | 8 (7.5) |
| Oral/pharyngeal/tonsillar | 10 (6.5) | 4 (3.7) | 4 (3.7) |
| Skin/body surface | 5 (3.2) | 1 (0.9) | 1 (0.9) |

Notes: Values are *n* (%) unless otherwise indicated; missing values are shown as *n*. Some patients contributed multiple specimens.
Abbreviations: Ct, cycle threshold; HIV, human immunodeficiency virus; IQR, interquartile range; MSM, men who have sex with men; qPCR, quantitative polymerase chain reaction; RPR, rapid plasma reagin.

### Adjusted qPCR Positivity and Mean Ct by Automated RPR and Syphilis Stage

Observed positivity was 74.3% (84/113 specimens) in primary syphilis and 61.0% (25/41 specimens) in secondary syphilis; positivity by RPR category within each stage is shown in Supplementary Table S3. Estimates of overall adjusted qPCR positivity remained between 65% and 68% across the range of automated RPR (Figure 2A and Table 2A). Stage-specific adjusted positivity (95% CrIs) was similar: 66.7% (58.7%–74.7%) in primary and 64.3% (46.9%–79.7%) in secondary syphilis. Within stages, however, the posterior median curves showed opposite trends across RPR values. Between RPR values of 1 and 100, adjusted positivity (95% CrIs) in primary syphilis decreased from 72.2% (60.9%–82.9%) to 61.2% (45.4%–75.1%), whereas that in secondary syphilis increased from 43.8% (16.7%–69.9%) to 82.9% (69.6%–91.1%). Consistent with these opposing trends, the secondary-to-primary ratio of the per-doubling aORs was 1.78 (95% CrI, 1.25–2.65), supporting a stage-by-RPR interaction (Table 2B).

**Figure 2.**
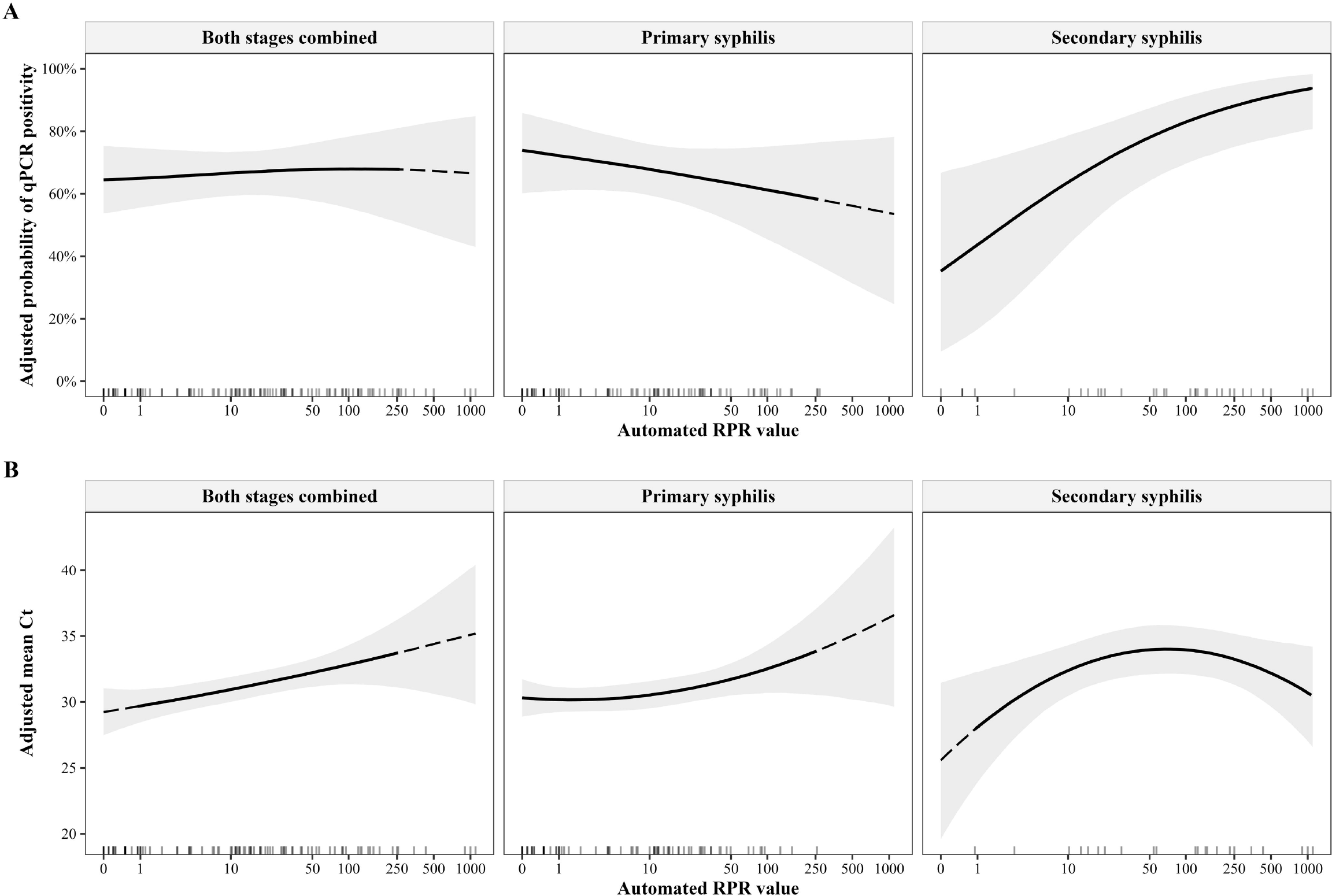
Adjusted *Treponema pallidum* qPCR Positivity and Mean Ct by Automated RPR and Syphilis Stage. Panel A shows adjusted qPCR positivity in 154 specimens; panel B shows adjusted mean Ct in 107 dual-target-positive specimens. Lines show posterior medians of estimates standardized over the respective analysis populations; bands show pointwise 95% CrIs. Combined-stage curves retain the observed stage composition. Solid segments cover observed patient-level RPR ranges, using the range common to both stages for combined-stage curves; dashed segments indicate extrapolation, and rug marks show observed patient-level RPR values. RPR is plotted on the log□(RPR+1) scale with tick labels showing the original values. Abbreviations: CrI, credible interval; Ct, cycle threshold; qPCR, quantitative polymerase chain reaction; RPR, rapid plasma reagin.

**Table 2.** Adjusted *Treponema pallidum* qPCR Positivity and Mean Ct by Automated RPR and Syphilis Stage.

**A. Adjusted qPCR positivity, % (95% CrI)**
| Automated RPR | Primary syphilis | Secondary syphilis | Both stages combined | Secondary – primary, percentage points |
| --- | --- | --- | --- | --- |
| Overall | 66.7 (58.7–74.7) | 64.3 (46.9–79.7) | — | –2.4 (–22.5 to 16.0) |
| 1 | 72.2 (60.9–82.9) | 43.8 (16.7–69.9) | 65.0 (55.6–74.6) | –28.6 (–57.7 to 0.6) |
| 10 | 67.9 (59.5–75.9) | 63.6 (43.6–78.6) | 66.6 (59.4–73.4) | –4.4 (–26.8 to 14.0) |
| 100 | 61.2 (45.4–75.1) | 82.9 (69.6–91.1) | 67.9 (55.4–78.3) | 21.3 (2.4 to 39.4) |

**B. Association of automated RPR with qPCR positivity: Adjusted ORs and ratio of ORs (95% CrI)**
| Measure | Primary syphilis | Secondary syphilis | Ratio of ORs (secondary/primary) |
| --- | --- | --- | --- |
| Per doubling of (RPR + 1) | 0.88 (0.70–1.11) | 1.57 (1.16–2.23) | 1.78 (1.25–2.65) |

**C. Adjusted mean Ct among dual-target-positive specimens, Ct (95% CrI)**
| Automated RPR | Primary syphilis | Secondary syphilis | Both stages combined |
| --- | --- | --- | --- |
| Overall | 31.24 (30.33–32.13) | 30.90 (28.79–33.01) | — |
| 1 | 30.18 (29.24–31.12) | 28.12 (23.95–32.29) | 29.71 (28.47–30.97) |
| 10 | 30.53 (29.52–31.58) | 32.38 (30.46–34.30) | 30.95 (30.02–31.91) |
| 100 | 32.52 (30.69–34.33) | 33.96 (32.11–35.73) | 32.84 (31.35–34.30) |

Notes: Analyses included 154 specimens for qPCR positivity and 107 for mean Ct. Values are posterior medians with 95% CrIs. Estimates in panels A and C were standardized over their respective analysis populations using the models described in the Methods. Overall estimates retain the analysis population’s RPR distribution, whereas combined-stage estimates retain its observed stage composition. In panel B, the secondary-to-primary ratio of adjusted ORs quantifies the stage-by-RPR interaction.
Abbreviations: CrI, credible interval; Ct, cycle threshold; OR, odds ratio; qPCR, quantitative polymerase chain reaction; RPR, rapid plasma reagin.

Among the 107 specimens included in the mean-Ct analysis, the median specimen-level mean Ct (IQR) was 30.20 (28.11–33.70) overall, 29.78 (27.77–32.89) in primary syphilis (83 specimens), and 32.82 (30.19–36.15) in secondary syphilis (24 specimens). The *TP0574* and *polA* Ct values were closely correlated (Pearson *r* = 0.966; Supplementary Figure S1). Overall, adjusted mean Ct increased by approximately 3 cycles between RPR values of 1 and 100 (Table 2C and Figure 2B). In primary syphilis, adjusted mean Ct was 31.24, and in secondary syphilis it was 30.90; the secondary-to-primary difference was −0.33 cycles (95% CrI, −2.69 to 2.07). Despite this similarity in level, the adjusted mean Ct curves across RPR differed in shape by stage (Figure 2B). In primary syphilis, adjusted mean Ct increased with RPR. In secondary syphilis, however, it increased to a peak near an RPR value of 68 and then decreased at higher RPR values, forming an inverted-U pattern.

### Predicted qPCR Positivity by Population Group

Secondary syphilis accounted for a higher proportion of specimens from MSM. Almost all specimens from non-MSM men were genitourinary, whereas oral or skin specimens were more common among MSM and women (Supplementary Table S4). Average predicted positivity (95% CrI) was highest for non-MSM men at 79.6% (72.2%–85.9%), compared with 55.4% (44.4%–66.0%) for MSM and 58.9% (36.1%–79.0%) for women. Among MSM, predicted positivity (95% CrI) increased from 36.8% (24.4%–55.0%) to 57.0% (43.8%–69.9%) as RPR value increased from 1 to 100, whereas it decreased slightly among non-MSM men and changed little among women (Figure 3).

**Figure 3.**
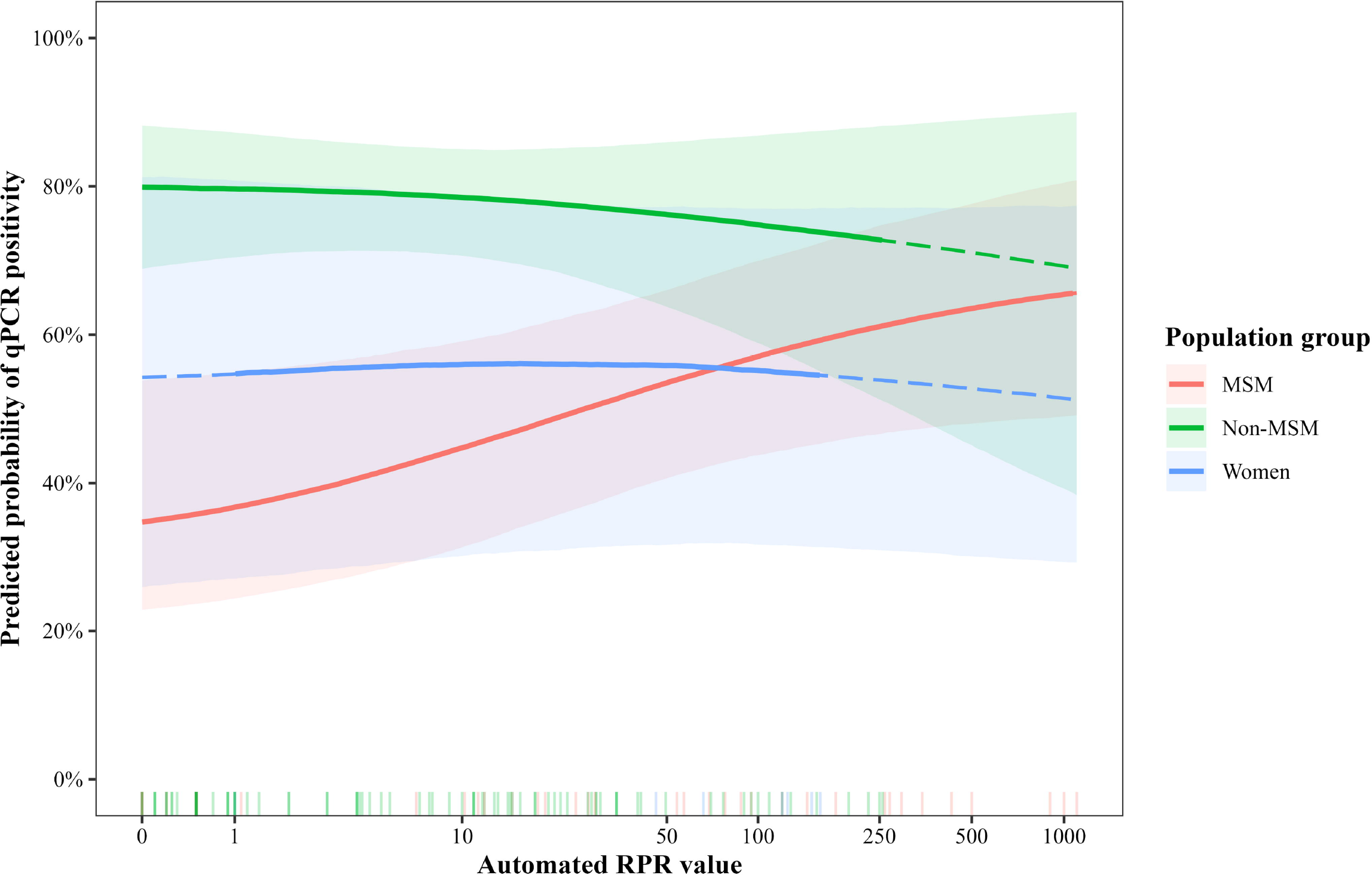
Predicted *Treponema pallidum* qPCR Positivity by Automated RPR and Population Group. Curves show posterior medians of predictions averaged within each population group; bands show pointwise 95% CrIs. Predictions retain each group’s covariate and facility composition (46 specimens from MSM, 96 from non-MSM men, and 12 from women). Solid segments cover each group’s observed patient-level RPR range, dashed segments indicate extrapolation, and rug marks show observed patient-level RPR values. RPR is plotted on the log□(RPR+1) scale with tick labels showing the original values. Abbreviations: CrI, credible interval; MSM, men who have sex with men; qPCR, quantitative polymerase chain reaction; RPR, rapid plasma reagin.

### Adjusted qPCR Positivity and Mean Ct by Specimen Collection Site

Adjusted qPCR positivity (95% CrI) was relatively high for genital/urethral/urinary (74.5%; 66.9%–81.4%) and anal/perianal/rectal specimens (74.5%; 46.0%–89.1%), and lower for oral/pharyngeal/tonsillar (36.6%; 11.1%–67.8%) and skin/body surface specimens (16.3%; 2.4%–54.7%) (Figure 4A; aORs in Supplementary Table S5A). In the observed data, 4 of 5 oral/pharyngeal/tonsillar specimens from patients with secondary syphilis were positive, whereas none of 5 specimens from patients with primary syphilis were positive. Adjusted mean Ct (95% CrI) ranged from 28.66 (25.41–31.94) for oral/pharyngeal/tonsillar to 31.34 (30.68–32.01) for genital/urethral/urinary specimens (Figure 4B; Ct contrasts in Supplementary Table S5B).

**Figure 4.**
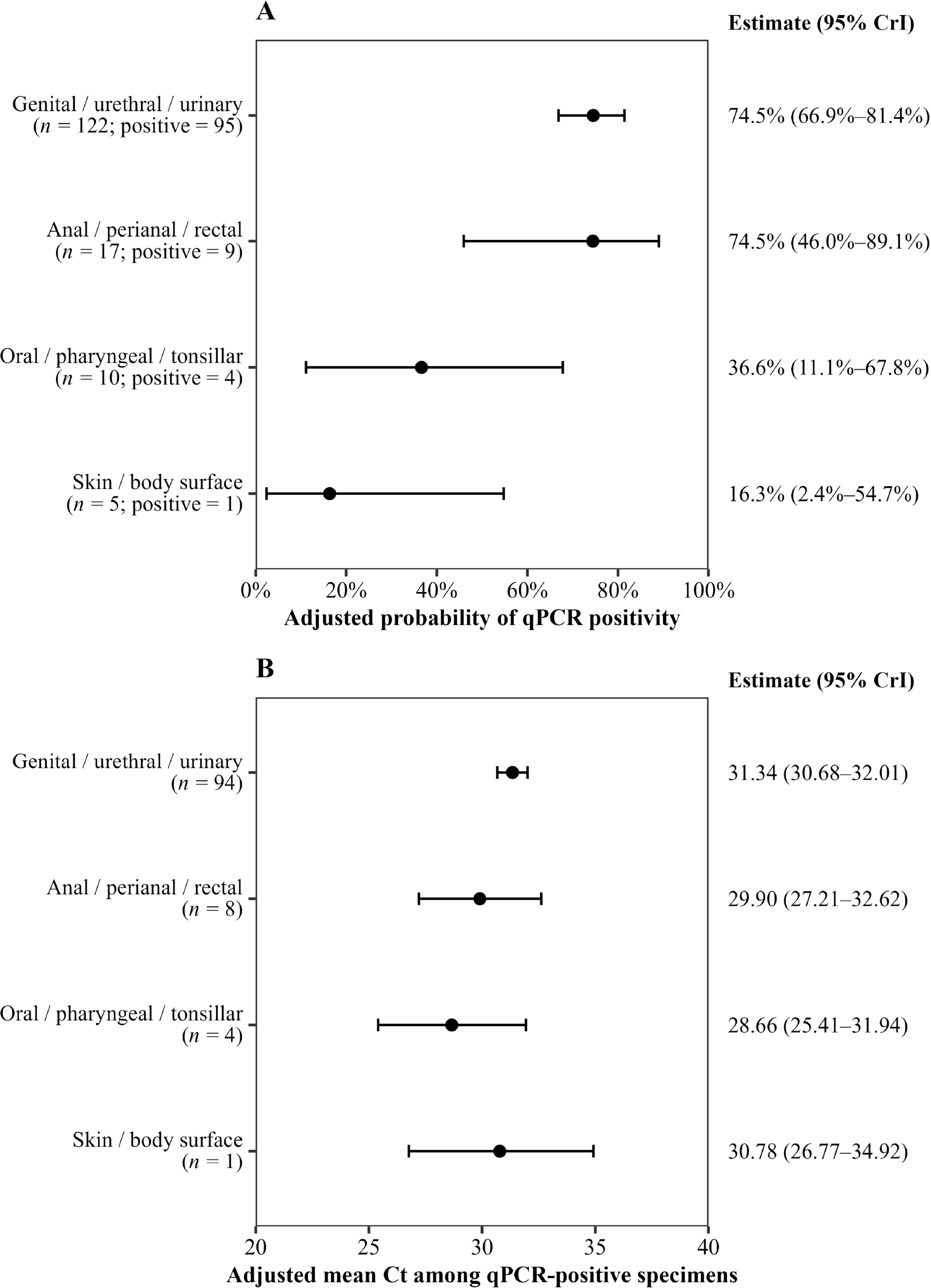
Adjusted *Treponema pallidum* qPCR Positivity and Mean Ct by Specimen Collection Site. Panel A shows adjusted qPCR positivity in 154 specimens; panel B shows adjusted mean Ct in 107 dual-target-positive specimens. Points show posterior medians of estimates standardized over the respective analysis populations; horizontal bars show 95% CrIs. Abbreviations: CrI, credible interval; Ct, cycle threshold; qPCR, quantitative polymerase chain reaction.

### Model Diagnostics and Sensitivity Analyses

All models converged, with no divergent transitions; posterior predictive checks showed no systematic lack of fit. Leave-one-out cross-validation favored a linear RPR specification for qPCR positivity and a quadratic specification for mean Ct (Supplementary Table S6). Across sensitivity analyses, the stage-specific qPCR aORs and their ratio and the stage-specific mean-Ct quadratic coefficients and their difference were consistent with the main analyses (Supplementary Table S7).

### qPCR Positivity in Blood Specimens

None of 131 blood specimens from 131 patients with early symptomatic syphilis were qPCR positive (95% CI, 0%–2.8%; Figure 1 and Supplementary Table S8).

## DISCUSSION

This multicenter study of specimens collected during routine care showed that adjusted qPCR positivity in primary syphilis was high even at low RPR and declined modestly as RPR rose. In secondary syphilis, by contrast, positivity rose progressively with RPR, indicating a stage-by-RPR interaction. Adjusted mean Ct increased with RPR in primary syphilis and followed an inverted-U pattern in secondary syphilis. In the separate blood analysis, qPCR yielded no positive results.

To our knowledge, these opposing stage-specific associations between RPR and qPCR positivity have not previously been described in nonblood specimens. A study of whole-blood specimens using RPR titer categories reported trends in opposite directions by stage [21]. On the other hand, studies that included nonblood specimens have generally combined stages when relating RPR to *T. pallidum* DNA detection and have reported inconsistent findings [6–8]. In our study, combined-stage positivity was nearly flat across RPR values, reflecting stage-specific trends that offset each other. Differences in stage distribution among studies may therefore contribute to this inconsistency.

In primary syphilis, the high qPCR positivity observed even at low RPR supports the potential value of qPCR when serologic findings do not yet establish the diagnosis. Early in infection, serologic confirmation may require repeat testing, which can lead to missed or delayed diagnoses when patients do not return [22]. qPCR of a nonblood specimen collected at the initial visit could provide direct evidence of infection without this delay, allowing timely treatment and reducing opportunities for transmission, although these outcomes were not evaluated in this study.

In the Ct analysis, adjusted mean values increased with RPR in primary syphilis. Primary lesions contain abundant DNA before RPR rises appreciably [22] and yield lower Ct than most secondary lesions [23]. As the systemic response to treponemal and host-derived lipoidal antigens develops [5, 24], treponemes are progressively cleared from the lesion, which may leave less DNA at higher RPR values. In secondary syphilis, however, adjusted mean Ct followed an inverted-U pattern across RPR values, a finding not described in earlier studies of Ct [7–9]. Secondary manifestations appear only after dissemination, when the host response is already established [5], so RPR values may not correspond to the DNA remaining at the lesion.

The reasons for the inverted-U pattern remain uncertain. Patients with secondary syphilis can also have primary manifestations [5], whose lower Ct could contribute to the pattern if the proportion of such specimens differed across RPR values. This could not be assessed because the type of manifestation at the collection site was insufficiently documented. The pattern also requires confirmation because estimates in this stage were based on few specimens, particularly at low RPR. The Ct findings also raise the question of whether the relationship between serologic activity and transmission potential differs by stage. Studies incorporating measures of organism viability or transmission are needed to determine whether transmission potential in secondary syphilis is lower at intermediate than at low or high RPR values [8, 9].

In population-group analyses, predicted positivity showed an upward trend with RPR among MSM, whereas its average level was highest for specimens from non-MSM men. These predictions describe each group’s observed profile rather than an independent population-group effect. Because only stage interacted with RPR in the model, the upward trajectory among MSM largely reflects their higher proportion of secondary syphilis. Differences in average positivity, by contrast, may partly reflect collection-site composition, as oral and skin specimens, which had the lowest adjusted positivity estimates, were more common among MSM and women than among non-MSM men.

Across collection sites, adjusted positivity was similar for genitourinary and anorectal specimens, consistent with the frequent detection previously reported for genital and anal lesion swabs [25]. By contrast, oral specimens had lower adjusted positivity. Secondary syphilis was the only stage with qPCR-positive oral specimens in our data, consistent with systematic oral-sampling studies reporting more frequent detection in secondary than in primary syphilis [6, 8, 9, 26]. Among the four positive oral specimens, the adjusted mean Ct was relatively low, compatible with reports of substantial treponemal DNA loads in saliva [26].

Blood qPCR offered no additional detection, as none of the 131 specimens was positive. Previous studies using different PCR methods have reported blood positivity of 3%–54% in primary and 15%–63% in secondary syphilis [8, 21, 25]. Here, collection-tube type was unavailable, and blood specimens were not centrifuged before DNA extraction. Differences in specimen processing and amplification methods may therefore explain the discrepancy.

A key strength of our study is the inclusion of specimens collected during routine care across Japan. Testing at a single central laboratory avoided interlaboratory variability in qPCR. The study also included non-MSM men and women, who have been underrepresented in earlier studies of *T. pallidum* detection. However, several limitations warrant consideration. First, specimen collection and automated RPR measurement followed local practice; observed differences in positivity and Ct may therefore reflect procedural as well as biological differences. Second, effects of the concurrent assay changes on positivity and Ct estimates could not be excluded. Third, population-group and collection-site estimates were subject to variation, and the granularity of the documentation limited our ability to fully break down analyses by manifestation and nonblood specimen types. Finally, participating facilities were not selected to be nationally representative.

In conclusion, our findings demonstrate high nonblood qPCR positivity in primary syphilis even at low or negative automated RPR values, where treponemal DNA recovery was also greatest. Rather than waiting for serologic maturation, these findings argue for integrating nonblood qPCR into the initial evaluation of suspected early syphilis to avert missed windows for intervention. While secondary syphilis profiles varied with RPR, prompt confirmation in either stage would optimize early patient management and curb onward transmission.

## Data Availability

All data produced in the present study are available upon reasonable request to the authors

## ACKNOWLEDGEMENTS

We thank the patients whose specimens and clinical information contributed to this study and the physicians and staff at the submitting institutions (listed in the Supplementary Material) for their assistance in submitting specimens and completing the case report forms.

## Author contributions

Conceptualization: A.K., Y.O., Y.A. Methodology: A.K., Y.O., Y.T., S.N., R.O., T.Y., Y.A. Investigation: Y.O., K.I., S.N., M.Y. Data curation: A.K., Y.O. Formal analysis: A.K. Resources: M.Y. Funding acquisition, project administration, and supervision: Y.A. Writing – original draft: A.K., Y.O. Writing – review & editing: all authors. All authors interpreted the findings, approved the final manuscript, and are accountable for the work.

## Data availability

Data are available from the corresponding author upon reasonable request.

