## Supplementary materials for "Stage-Specific Associations of Automated RPR With *Treponema pallidum* qPCR Positivity and Cycle Threshold in Early Symptomatic Syphilis: A Multicenter Study Using Specimens Collected During Routine Care"

A Multicenter Study Using Specimens Collected During Routine Care

**Supplementary Material**

Supplementary Methods, Supplementary Figure S1, and Supplementary Tables S1–S8,

and Specimen-submitting institutions

**Supplementary Methods**

**Model specification**

The qPCR-positivity and mean-Ct models were fitted separately at the specimen level using Bayesian logistic and normal regression, respectively.

For each outcome model, let $j=1, \ldots, J$ index patients and $n=1, \ldots, N$ index specimens included in that analysis. Let $j\left( n \right)$ denote the patient contributing specimen $n$, with $j=j\left( n \right)$ in specimen-level equations. Let $k\left( n \right)$ denote the specimen's collection-site category and $f\left( j\left( n \right) \right)$ the patient's facility. Collection-site categories were genital/urethral/urinary, anal/perianal/rectal, oral/pharyngeal/tonsillar, and skin/body surface. Genital/urethral/urinary was the reference category in both outcome models. Let $S_{j}$, $F_{j}$, $H_{j}$, and $P_{j}$ equal 1 for secondary syphilis, female sex assigned at birth, HIV infection, and previous syphilis, respectively, and 0 otherwise. Population group was not included as a covariate in either outcome model. Let $A_{j}$ denote age in years and $\mathrm{RPR}_{j}$ the automated RPR value. Age was centered at 40 years and scaled in 10-year units as $A_{j}^{*}=\frac{A_{j} - 40}{10}$. Negative RPR results were coded as 0, and values below the reporting limit were assigned half that limit. Automated RPR was transformed as $X_{j}=\log_{2} \left( \mathrm{RPR}_{j}+1 \right)$, so that a 1-unit increase in $X_{j}$ corresponded to a doubling of $\left( \mathrm{RPR}_{j}+1 \right)$.

In the outcome-model equations, $\alpha$ denotes the intercept, $\beta$ the fixed-effect coefficients, and $b$ the facility-specific random intercept. The first subscript identifies the outcome model ($Y$, qPCR positivity; $C$, mean Ct), and the remaining subscripts identify the corresponding variable, term, or facility. Throughout, $\mathrm{Normal}\left( \mu, \sigma\right)$ denotes a Normal distribution with mean $\mu$ and standard deviation (SD) $\sigma$, and $\mathrm{half}\text{-}\mathrm{Normal}\left( s \right)$ denotes the distribution obtained by restricting a zero-mean Normal distribution with SD (s) to nonnegative values.

For qPCR positivity, let $Y_{n}=1$ for a positive result and 0 otherwise, and let $p_{n}$ denote the modeled probability of a positive result for specimen $n$. We modeled $Y_{n}\sim\mathrm{Bernoulli}\left( p_{n} \right),$ with a linear association between transformed RPR and the log odds of qPCR positivity within each stage. A stage-by-RPR interaction allowed the slope to differ between stages, giving the following linear predictor:

$$\begin{aligned} \mathrm{logit}\left( p_{n} \right)=\alpha_{Y}+\beta_{Y,\mathrm{site},k\left( n \right)}+\beta_{Y,S}S_{j}+\beta_{Y,X}X_{j}+\beta_{Y,SX}S_{j}X_{j}+\beta_{Y, A}A_{j}^{*}+\beta_{Y,F}F_{j}+\beta_{Y, H}H_{j}+\beta_{Y,P}P_{j}+b_{Y,f\left( j\left( n \right) \right)}. \end{aligned}$$

The facility-specific random intercepts $b_{Y,f}$ followed independent $\mathrm{Normal}\left( 0,\tau_{Y} \right)$ distributions, where $\tau_{Y}$ denotes the between-facility standard deviation; a patient-level random intercept was not included because repeated specimens from the same patient were uncommon. The stage-by-RPR interaction was quantified as $\exp\left( \beta_{Y,SX} \right)$, the adjusted odds ratio per doubling of $\left( \mathrm{RPR}+1 \right)$ in secondary syphilis divided by the corresponding adjusted odds ratio in primary syphilis.

For the mean-Ct model, $C_{n}$ was the mean of the *TP0574* (TpN47) and *polA* Ct values. $x_{j}=X_{j}-c_{C}$, where $c_{C}=3.65517$corresponded to an automated RPR of 11.6. In the equation below, $\beta_{C,L}$ is the linear coefficient in primary syphilis, and $\beta_{C,LS}$ is the secondary-minus-primary difference in that coefficient. The quadratic coefficients are $\beta_{C,Q1}$ for primary syphilis and $\beta_{C,Q2}$ for secondary syphilis. The model was

$$C_{n}\sim\mathrm{Normal}\left( \mu_{C,n},\sigma_{C} \right),$$

$$\begin{aligned} \mu_{C,n}=\alpha_{C}+\beta_{C,\mathrm{site},k\left( n \right)}+\beta_{C,S}S_{j}+\left( \beta_{C,L}+\beta_{C,LS}S_{j} \right)x_{j}+\left[ \left( 1-S_{j} \right)\beta_{C,Q1}+S_{j}\beta_{C,Q2} \right]x_{j}^{2}+\beta_{C,A}A_{j}^{*}+\beta_{C,F}F_{j}+\beta_{C,H}H_{j}+\beta_{C,P}P_{j}+b_{C,f\left( j\left( n \right) \right)}. \end{aligned}$$

The facility-specific random intercepts $b_{C,f}$ followed independent $\mathrm{Normal}\left( 0,\tau_{C} \right)$ distributions, with $\tau_{C}$ denoting the between-facility standard deviation. The between-stage contrast in the quadratic RPR coefficient reported in Supplementary Table S7B was $\beta_{C,Q2}-\beta_{C,Q1}$.

**Missing age and automated RPR values**

For each outcome, the specimen-level model was fitted jointly with patient-level submodels for age and transformed automated RPR ($X_{j}$). Both submodels contributed likelihood terms for every patient included in that analysis, using recorded values as observed data and treating missing values as latent variables. The age submodel was

$$A_{j}\sim\mathrm{Normal}\left( \mu_{A,j}, \sigma_{A} \right) truncated to \left[ 18, 100 \right],$$

$$\mu_{A,j}=\alpha_{A}+\gamma_{A,S}S_{j}+\gamma_{A,F}F_{j}+\gamma_{A,H}H_{j}+\gamma_{A,P}P_{j}+b_{A,f\left( j \right)}.$$

The transformed-RPR submodel was

$$X_{j}\sim\mathrm{Normal}\left( \mu_{X,j}, \sigma_{X} \right) truncated to \left[ 0, \infty\right),$$

$$\mu_{X,j}=\alpha_{X}+\gamma_{X,S}S_{j}+\gamma_{X,A}A_{j}^{*}+\gamma_{X,F}F_{j}+\gamma_{X,H}H_{j}+\gamma_{X,P}P_{j}+b_{X,f\left( j \right)}.$$

Here, $\gamma$ denotes the fixed-effect coefficients in the age and transformed-RPR submodels. The facility-specific random intercepts were specified as $b_{A,f}\overset{iid}{\sim}\mathrm{Normal}\left( 0, \tau_{A} \right)$ and $b_{X,f}\overset{iid}{\sim}\mathrm{Normal}\left( 0, \tau_{X} \right)$, where $\tau_{A}$and $\tau_{X}$denote the corresponding between-facility SDs. Within each fit, the facility effects for the outcome, age, and RPR components were mutually independent *a priori*. Missingness of age and RPR was assumed ignorable conditional on the observed data in the joint model.

**Prior distributions**

In the qPCR outcome component, the intercept and all estimated fixed-effect coefficients had $\mathrm{Normal}\left( 0, 2.5 \right)$ priors, and $\tau_{Y}$ had a$\mathrm{half}\text{-}\mathrm{Normal}\left( 1 \right)$ prior.

In the mean-Ct component, $\alpha_{C}$ had a $\mathrm{Normal}\left( 30, 8 \right)$ prior, the stage-specific quadratic coefficients $\beta_{C,Q1}$ and $\beta_{C,Q2}$ had $\mathrm{Normal}\left( 0, 0.20 \right)$ priors, and all other estimated fixed-effect coefficients had $\mathrm{Normal}\left( 0, 2.5 \right)$ priors. The parameters $\sigma_{C}$ and $\tau_{C}$ had $\mathrm{half}\text{-}\mathrm{Normal}\left( 5 \right)$ and $\mathrm{half}\text{-}\mathrm{Normal}\left( 1 \right)$ priors, respectively.

For the age submodel, $\alpha_{A}$ had a $\mathrm{Normal}\left( 40, 15 \right)$ prior, each $\gamma$ coefficient had a $\mathrm{Normal}\left( 0, 10 \right)$ prior, and $\sigma_{A}$ and $\tau_{A}$ had $\mathrm{half}\text{-}\mathrm{Normal}\left( 15 \right)$ and $\mathrm{half}\text{-}\mathrm{Normal}\left( 10 \right)$ priors, respectively. For the transformed-RPR submodel, $\alpha_{X}$ had a $\mathrm{Normal}\left( 3, 3 \right)$ prior, each $\gamma$ coefficient had a $\mathrm{Normal}\left( 0, 2 \right)$ prior, and $\sigma_{X}$and $\tau_{X}$had $\mathrm{half}\text{-}\mathrm{Normal}\left( 2 \right)$ and $\mathrm{half}\text{-}\mathrm{Normal}\left( 1.5 \right)$ priors, respectively.

**Standardized predictions**

For each posterior draw, covariates were retained at their observed values or, if missing, their sampled values unless set as specified below; fitted facility effects were retained. For overall adjusted estimates by stage or collection site, only the corresponding variable was set. Model-based probabilities or mean Ct values were then averaged over all specimens included in that analysis.

For RPR curves, the same calculation and averaging were repeated with RPR set to each grid value, retaining each specimen’s observed stage for combined-stage curves and setting stage to primary or secondary syphilis for stage-specific curves. Both outcome models used a common grid from 0 to 1100, the maximum automated RPR observed among patients included in the main qPCR analysis, with equal spacing on the $\log_{2} \left( \mathrm{RPR}+1 \right)$ scale.

Population-group qPCR predictions used the same calculations, but probabilities were averaged only over specimen records within each group. For group-specific curves, RPR was set to each grid value.

**Posterior computation and model checking**

Posterior distributions were sampled using Stan’s No-U-Turn sampler. Each model was run with 4 chains of 4000 iterations, including 2000 warm-up iterations per chain, yielding 8000 post-warm-up draws in total. Across models, the target acceptance probability was set between 0.95 and 0.99, and the maximum tree depth was 15. Across all reported fits, the maximum $\hat{R}$ was 1.003 and the minimum bulk effective sample size was 1629. No post-warm-up transitions were divergent, and none reached the maximum tree depth.

For qPCR positivity, posterior predictive checks compared observed and replicated counts of positive specimens overall and positivity proportions within strata defined by stage and collection site. For mean Ct, checks compared observed and replicated distributional summaries overall and by stage. Within each stage, observed and replicated means and SDs were also compared for the observed RPR categories (<10, 10 to <100, and ≥100).

**Alternative RPR functional forms**

We compared the main linear RPR specification for qPCR positivity with a quadratic alternative, and the main quadratic specification for mean Ct with a linear alternative.

For the alternative quadratic qPCR model only, we defined $x_{j}=X_{j}-3.70044$, where 3.70044 was the median $X_{j}$ among patients included in the qPCR analysis who had recorded RPR values (corresponding to an automated RPR of 12.0). The RPR terms in the main qPCR model were replaced by $\theta_{L}x_{j}+\theta_{Q}{x_{j}}^{2}+\theta_{LS}S_{j}x_{j}+\theta_{QS}S_{j}{x_{j}}^{2}.$ The linear and stage-by-linear coefficients $\theta_{L}$ and $\theta_{LS}$ each had a $\mathrm{Normal}\left( 0, 2.5 \right)$ prior, whereas the quadratic and stage-by-quadratic coefficients $\theta_{Q}$ and $\theta_{QS}$ each had a $\mathrm{Normal}\left( 0, 0.10 \right)$ prior. All other model terms and priors were unchanged.

For mean Ct, the alternative linear model used the uncentered terms $X_{j}$ and $S_{j}X_{j}$, omitted the two stage-specific quadratic terms, and otherwise retained the main mean-Ct model specification.

For each outcome, the main and alternative models were compared using Pareto-smoothed importance-sampling leave-one-specimen-out cross-validation. The comparison used specimen-level outcome log-likelihood contributions, excluding contributions from the age and RPR submodels. Moment matching was applied to the alternative quadratic qPCR model.

**Sensitivity analysis**

For both outcome models, we conducted sensitivity analyses with one change at a time: excluding patients with previous syphilis, restricting the analysis to specimens from patients with recorded age and automated RPR values, using narrower Normal priors for the outcome-model coefficients, or defining the outcomes using only *polA,* whose primers were unchanged. In the narrower-prior analyses, the prior SD was reduced from 2.5 to 1.5 for the qPCR outcome-model intercept and all estimated fixed-effect coefficients, and for the mean-Ct fixed-effect coefficients other than the intercept and the two stage-specific quadratic coefficients.

For qPCR positivity, polA detection was defined by a recorded Ct, with 152 specimens included after excluding 2 without a documented result.　An additional analysis excluded the 8 patients whose recorded diagnosis may have depended on qPCR.

For mean Ct, the *polA* Ct analysis used the same 107 specimens as the main mean-Ct analysis. A separate prior-sensitivity analysis reduced the prior SD for both stage-specific quadratic coefficients from 0.20 to 0.10.

**Blood Specimen Processing**

Blood specimens were submitted in either EDTA-containing tubes or serum-separator tubes containing a clot activator. Specimens were not centrifuged before DNA extraction and were analyzed as submitted. Specimen-level information on collection-tube type was unavailable, and blood specimens were not further classified by blood fraction.

**Supplementary Figure**

**Supplementary Figure S1. Correlation Between *TP0574* and *polA* Ct Values Among Dual-Target-Positive Specimens**

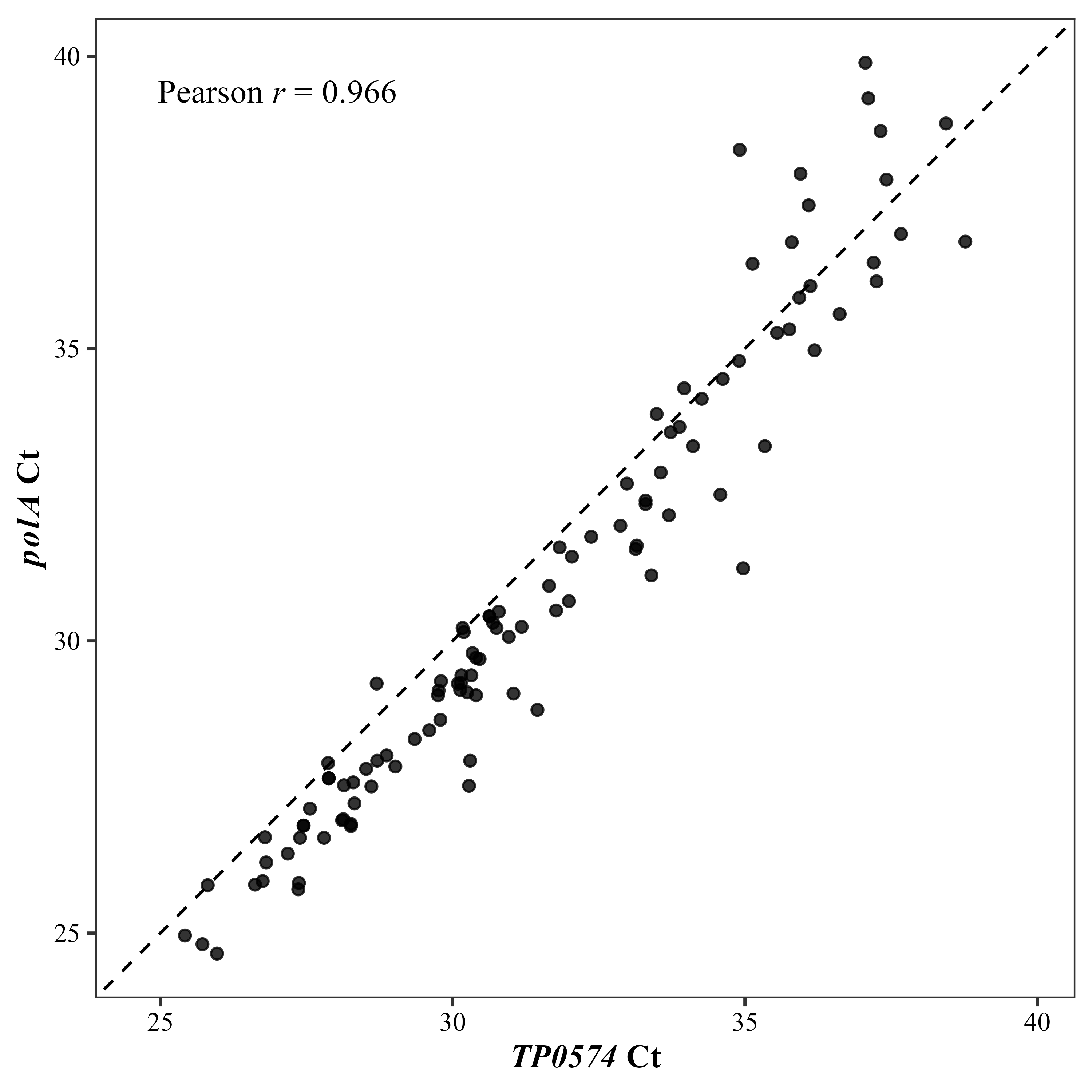

Notes: Each point represents 1 of 107 specimens with numeric Ct values for both qPCR targets, *TP0574* and *polA*. The dashed diagonal is the line of identity; the displayed coefficient is Pearson *r*. The mean difference between targets was 0.62 cycles (*TP0574* － *polA*).

Abbreviations: Ct, cycle threshold; qPCR, quantitative polymerase chain reaction.

**Supplementary Tables**

**Supplementary Table S1. Patient and Specimen Characteristics by Syphilis Stage**

| Characteristic | Overall | Primary syphilis | Secondary syphilis |
| --- | --- | --- | --- |
| **A. Patient-level characteristics** | | | |
| Patients, No. | 145 | 108 | 37 |
| Age, y, median (IQR) | 41.0 (32.5–51.0) | 41.0 (32.2–51.8) | 38.0 (34.0–48.0) |
| Missing, No. | 6 | 6 | 0 |
| Population group, No. (%) |  |  |  |
| MSM | 41 (28.3) | 18 (16.7) | 23 (62.2) |
| Non-MSM men | 95 (65.5) | 84 (77.8) | 11 (29.7) |
| Women | 9 (6.2) | 6 (5.6) | 3 (8.1) |
| Documented HIV infection, No. (%) | 22 (15.2) | 7 (6.5) | 15 (40.5) |
| Previous syphilis, No. (%) | 31 (21.4) | 14 (13.0) | 17 (45.9) |
| Missing, No. | 2 | 0 | 2 |
| Automated RPR value, median (IQR) | 12.0 (0.9–63.8) | 7.3 (0.5–28.2) | 88.9 (15.8–222.1) |
| Negative, No. (%) | 7 (4.8) | 6 (5.6) | 1 (2.7) |
| Missing, No. | 11 | 8 | 3 |
| **B. Specimen-level characteristics** | | | |
| Specimens, No. | 154 | 113 | 41 |
| Collection site, No. (%) |  |  |  |
| Genital/urethral/urinary | 122 (79.2) | 105 (92.9) | 17 (41.5) |
| Anal/perianal/rectal | 17 (11.0) | 3 (2.7) | 14 (34.1) |
| Oral/pharyngeal/tonsillar | 10 (6.5) | 5 (4.4) | 5 (12.2) |
| Skin/body surface | 5 (3.2) | 0 (0.0) | 5 (12.2) |

Notes: Main nonblood qPCR analysis cohort. Percentages use patient totals in panel A and specimen totals in panel B. Age and RPR summaries exclude missing values; RPR coding is described in Supplementary Methods.

Abbreviations: HIV, human immunodeficiency virus; IQR, interquartile range; MSM, men who have sex with men; qPCR, quantitative polymerase chain reaction; RPR, rapid plasma reagin.

**Supplementary Table S2. Numbers of Nonblood Specimens by qPCR Assay Configuration**

| Analysis | Original configuration | Updated configuration | Total |
| --- | --- | --- | --- |
| qPCR positivity | 127 | 27 | 154 |
| Mean Ct | 91 | 16 | 107 |

Notes: Specimens included in the mean Ct analysis were a subset of those included in the qPCR-positivity analysis.

Abbreviations: Ct, cycle threshold; qPCR, quantitative polymerase chain reaction.

**Supplementary Table S3. Observed *Treponema pallidum* qPCR Positivity by Syphilis Stage and Automated RPR Category**

| Syphilis Stage | Automated RPR category | Specimens, No. | qPCR positive, No. (%) |
| --- | --- | --- | --- |
| Primary syphilis |  |  |  |
|  | Overall | 113 | 84 (74.3%) |
|  | <1 | 29 | 25 (86.2%) |
|  | 1 to <10 | 26 | 18 (69.2%) |
|  | 10 to <50 | 32 | 29 (90.6%) |
|  | 50 to <100 | 7 | 5 (71.4%) |
|  | 100 to <250 | 6 | 2 (33.3%) |
|  | ≥250 | 4 | 1 (25.0%) |
| Secondary syphilis |  |  |  |
|  | Overall | 41 | 25 (61.0%) |
|  | <1 | 6 | 1 (16.7%) |
|  | 1 to <10 | 1 | 1 (100.0%) |
|  | 10 to <50 | 7 | 6 (85.7%) |
|  | 50 to <100 | 4 | 3 (75.0%) |
|  | 100 to <250 | 10 | 6 (60.0%) |
|  | ≥250 | 9 | 7 (77.8%) |

Notes: Category rows exclude specimens with missing observed automated RPR.

Abbreviations: qPCR, quantitative polymerase chain reaction; RPR, rapid plasma reagin.

**Supplementary Table S4. Syphilis Stage and Specimen Collection Site by Population Group**

| Characteristic | | MSM | Non-MSM | Women |
| --- | --- | --- | --- | --- |
| Patients, No. | | 41 | 95 | 9 |
| Specimens, No. | | 46 | 96 | 12 |
| Syphilis stage, No. (%) | |  |  |  |
|  | Primary | 19 (41.3) | 85 (88.5) | 9 (75.0) |
|  | Secondary | 27 (58.7) | 11 (11.5) | 3 (25.0) |
| Specimen collection site, No. (%) | |  |  |  |
|  | Genital/urethral/urinary | 19 (41.3) | 94 (97.9) | 9 (75.0) |
|  | Anal/perianal/rectal | 17 (37.0) | 0 | 0 |
|  | Oral/pharyngeal/tonsillar | 7 (15.2) | 0 | 3 (25.0) |
|  | Skin/body surface | 3 (6.5) | 2 (2.1) | 0 |

Notes: Values are n (%) of specimens unless otherwise indicated. Some patients contributed more than 1 specimen.

Abbreviation: MSM, men who have sex with men.

**Supplementary Table S5. Additional Posterior Estimates From Models of *Treponema pallidum* qPCR Positivity and Mean Ct**

A. qPCR positivity model

| Model component | | Effect measure or unit | Posterior median | 95% CrI |
| --- | --- | --- | --- | --- |
| Collection site | |  |  |  |
|  | genital/urethral/urinary | Reference | — | — |
|  | anal/perianal/rectal | Adjusted OR vs genital/urethral/urinary | 0.99 | 0.17–5.52 |
|  | oral/pharyngeal/tonsillar | Adjusted OR vs genital/urethral/urinary | 0.12 | 0.01–0.75 |
|  | skin/body surface | Adjusted OR vs genital/urethral/urinary | 0.03 | 0.002–0.34 |
| Facility random-intercept SD | | Log-odds scale | 0.83 | 0.07–1.93 |

B. Mean Ct model

| Model component | | Effect measure or unit | Posterior median | 95% CrI |
| --- | --- | --- | --- | --- |
| Stage term: secondary vs primary syphilis | | Adjusted mean difference in Ct, cycles | 2.01 | −0.04 to 4.11 |
| RPR association within primary syphilis | | Adjusted mean difference in Ct per doubling of (RPR + 1), cycles | 0.39 | 0.02–0.74 |
| Difference in the RPR association: secondary − primary | | Difference in the above, cycles | 0.73 | −0.23 to 1.70 |
| Collection site: | |  |  |  |
|  | genital/urethral/urinary | Reference | — | — |
|  | anal/perianal/rectal | Adjusted mean difference vs genital/urethral/urinary, cycles | −1.45 | −4.28 to 1.45 |
|  | oral/pharyngeal/tonsillar | Adjusted mean difference vs genital/urethral/urinary, cycles | −2.69 | −6.03 to 0.74 |
|  | skin/body surface | Adjusted mean difference vs genital/urethral/urinary, cycles | −0.57 | −4.59 to 3.60 |
| Residual SD | | Ct scale | 3.11 | 2.69–3.65 |
| Facility random-intercept SD | | Ct scale | 1.04 | 0.07–2.32 |

Notes: Models are described in the Supplementary Methods. In panel B, the stage contrast and local RPR slope terms on the $\log_{2} \left( RPR+1 \right)$ scale are evaluated at an automated RPR of 11.6.

Abbreviations: CrI, credible interval; Ct, cycle threshold; OR, odds ratio; qPCR, quantitative polymerase chain reaction; RPR, rapid plasma reagin; SD, standard deviation.

**Supplementary Table S6. Comparison of Functional Forms for Automated RPR Using Pareto-Smoothed Importance-Sampling Leave-One-Out Cross-Validation**

| Model | | ELPD difference | SE of difference | Maximum Pareto *k* |
| --- | --- | --- | --- | --- |
| qPCR positivity | | | | |
|  | Main linear model | Reference | — | 0.61 |
|  | Alternative quadratic model | −1.33 | 1.29 | 0.68 |
| Mean Ct | | | | |
|  | Main quadratic model | Reference | — | 0.68 |
|  | Alternative linear model | −3.88 | 2.02 | 0.59 |

Notes: ELPD differences are relative to the main analysis model for the same outcome; a negative value indicates worse expected predictive performance than the main model.

Abbreviations: Ct, cycle threshold; ELPD, expected log predictive density; qPCR, quantitative polymerase chain reaction; SE, standard error.

**Supplementary Table S7. Sensitivity Analyses for the *Treponema pallidum* qPCR Positivity and Mean Ct Models**

A. qPCR positivity: adjusted odds ratios per 1-unit increase in $\log_{2} \left( RPR+1 \right)$, median (95% CrI)

| Analysis | Specimens, No. | Patients, No. | Primary syphilis | Secondary syphilis | Ratio, secondary / primary |
| --- | --- | --- | --- | --- | --- |
| Main model | 154 | 145 | 0.88 (0.70–1.11) | 1.57 (1.16–2.23) | 1.78 (1.25–2.65) |
| Age-and-RPR complete cases | 140 | 133 | 0.89 (0.70–1.12) | 1.60 (1.18–2.30) | 1.81 (1.26–2.74) |
| Exclusion of patients with previous syphilis | 119 | 114 | 0.83 (0.62–1.08) | 1.36 (0.78–2.43) | 1.63 (0.93–3.03) |
| Fixed-effect prior SD 1.5 | 154 | 145 | 0.91 (0.73–1.12) | 1.39 (1.06–1.87) | 1.54 (1.13–2.12) |
| Exclusion of 8 patients with possible diagnostic ascertainment | 145 | 137 | 0.92 (0.73–1.17) | 1.61 (1.17–2.29) | 1.74 (1.21–2.58) |
| *polA* detection as the outcome | 152 | 143 | 0.90 (0.72–1.13) | 1.57 (1.15–2.22) | 1.74 (1.22–2.59) |

B. Mean Ct: quadratic coefficient on centered $\log_{2} \left( RPR+1 \right)$, median (95% CrI)

| Analysis | Specimens, No. | Patients, No. | Primary syphilis | Secondary syphilis | Between-stage difference,  secondary − primary |
| --- | --- | --- | --- | --- | --- |
| Main dual-target mean Ct model | 107 | 105 | 0.084 (−0.068 to 0.229) | −0.226 (−0.412 to −0.040) | −0.310 (−0.545 to −0.071) |
| Age-and-RPR complete cases | 101 | 99 | 0.072 (−0.079 to 0.217) | −0.228 (−0.421 to −0.032) | −0.297 (−0.539 to −0.050) |
| Exclusion of patients with previous syphilis | 91 | 90 | 0.077 (−0.072 to 0.231) | −0.170 (−0.433 to 0.101) | −0.250 (−0.546 to 0.054) |
| Fixed-effect prior SD 1.5 | 107 | 105 | 0.067 (−0.081 to 0.214) | −0.210 (−0.387 to −0.036) | −0.278 (−0.505 to −0.044) |
| Quadratic-term prior SD 0.10 | 107 | 105 | 0.058 (−0.068 to 0.180) | −0.137 (−0.274 to 0.006) | −0.194 (−0.384 to −0.001) |
| *polA* Ct as the outcome | 107 | 105 | 0.102 (−0.052 to 0.259) | −0.241 (−0.428 to −0.053) | −0.343 (−0.580 to −0.097) |

Notes: Complete-case analyses included only specimens from patients with recorded age and automated RPR. The diagnostic-ascertainment analysis excluded the 8 patients whose recorded diagnosis may have depended on qPCR. The *polA* Ct analysis used *polA* Ct in the same 107 specimens as the main mean-Ct analysis. Full specifications are provided in the Supplementary Methods.

Abbreviations: CrI, credible interval; Ct, cycle threshold; qPCR, quantitative polymerase chain reaction; RPR, rapid plasma reagin; SD, standard deviation.

**Supplementary Table S8. Blood *Treponema Pallidum* qPCR Positivity in Blood Specimens From Patients With Early Symptomatic Syphilis**

| Syphilis stage | Blood specimens tested, No. | qPCR-positive specimens, No. (%) | 95% CI, % |
| --- | --- | --- | --- |
| Early symptomatic syphilis | 131 | 0 (0.0) | 0.0–2.8 |
| Primary syphilis | 101 | 0 (0.0) | 0.0–3.6 |
| Secondary syphilis | 30 | 0 (0.0) | 0.0–11.6 |

Notes: Exact 95% CIs were calculated by the 2-sided Clopper–Pearson method.

Abbreviations: CI, confidence interval; qPCR, quantitative polymerase chain reaction.

**Specimen-submitting institutions**

Anjo Kosei Hospital (Aichi)

Department of Legal Medicine, Kyorin University School of Medicine (Tokyo)

Fukuoka Wajiro Hospital (Fukuoka)

Hashimoto Municipal Hospital (Wakayama)

Hiratsuka City Hospital (Kanagawa)

Iguchi Urological Clinic, Kameari (Tokyo)

Itabashi Chuo Medical Center (Tokyo)

Jo Urology Clinic (Okayama)

KARADA Internal Medicine Clinic, Gotanda (Tokyo)

Matsuki Urological Clinic (Kagawa)

Matsunami General Hospital (Gifu)

National Hospital Organization Nishi-Saitama Chuo Hospital (Saitama)

National Hospital Organization Shinshu Ueda Medical Center (Nagano)

Nishifunabashi Urological Clinic (Chiba)

Nomura Clinic, Nipponbashi (Osaka)

Nomura Clinic, Umeda (Osaka)

Osaka Women’s and Children’s Hospital (Osaka)

Saga University Hospital (Saga)

Seo Clinic (Hiroshima)

Shirakaba Clinic (Tokyo)

Takayama Urological Tsukinoura Clinic (Fukuoka)

The Jikei University Hospital (Tokyo)

Tokyo Metropolitan Cancer and Infectious Diseases Center Komagome Hospital (Tokyo)

Tokyo Metropolitan Toshima Hospital (Tokyo)

Toneri Urology Clinic (Tokyo)

Toyama City Hospital (Toyama)

Yoh Urology and Dermatology Clinic (Aichi)

Yokoyama Urological Clinic (Okayama)
